# Doctors of chiropractic working with or within integrated health care delivery systems: a scoping review protocol

**DOI:** 10.1101/2020.08.11.20170399

**Authors:** Eric J. Roseen, Aisha B. Kasali, Kelsey Corcoran, Kelsey Masselli, Lance Laird, Robert Saper, Daniel P. Alford, Ezra Cohen, Anthony Lisi, Steven J. Atlas, Jonathan F. Bean, Roni Evans, André Bussières

**Affiliations:** Department of Family Medicine, Boston University School of Medicine and Boston Medical Center, Boston, MA, USA; Department of Rehabilitation Sciences, Massachusetts General Hospital Institute for Health Professions, Boston, MA, USA; New England Geriatric Research Education and Clinical Center, Boston Veterans Affairs Healthcare System, Boston, MA, USA; Pain Research, Informatics, Multimorbidities and Education (PRIME) Center, VA Connecticut Healthcare System, West Haven, CT, USA; Yale School of Medicine, Yale University, New Haven, CT, USA; Clinical Addiction Research and Education Unit, Section of General Internal Medicine, Boston University School of Medicine and Boston Medical Center, Boston, MA, USA; Department of Pediatrics, Boston University School of Medicine and Boston Medical Center, Boston, MA, USA; Division of General Internal Medicine, Massachusetts General Hospital, Boston, MA; Harvard Medical School, Boston, MA, USA; Spaulding Rehabilitation Hospital, Boston, MA; Integrative Health & Wellbeing Research Program, Earl E. Bakken Center for Spirituality and Healing, University of Minnesota, Minneapolis, MN, USA; Département Chiropratique, Université du Québec à Trois-Rivières, Trois-Rivières, Québec, Canada; School of Physical and Occupational Therapy, Faculty of Medicine, McGill University, Montreal, Québec, Canada

## Abstract

**Introduction:** Back and neck pain are the leading causes of disability worldwide. Doctors of chiropractic (DCs) are trained to manage these common conditions and can provide nonpharmacologic treatment aligned with international clinical practice guidelines. Although DCs practice in at least 90 countries, chiropractic care is often not available within integrated health care delivery systems. A lack of DCs in private practice, particularly in low-income communities, may also limit access to chiropractic care. Improved collaboration between medical providers and community-based DCs, or embedding DCs in medical settings such as hospitals or community health centers, will improve access to evidence-based care for musculoskeletal conditions.

**Methods and analyses:** This scoping review will map studies of DCs working with or within integrated health care delivery systems. We will use the recommended six-step approach for scoping reviews. We will search three electronic data bases including Medline, Embase, and Web of Science. Two investigators will independently review all titles and abstracts to identify relevant records, screen the full-text articles of potentially admissible records, and systematically extract data from selected articles. We will include studies published in English from 1998 to 2020 describing medical settings that have established formal relationships with community-based DCs (e.g., shared medical record) or where DCs practice in medical settings. Data extraction and reporting will be guided by the Proctor Conceptual Model for Implementation Research, which has three domains: clinical intervention; implementation strategies; and outcome measurement. Stakeholders from diverse clinical fields will offer feedback on the implications of our findings via a web-based survey.

**Ethics and dissemination:** Ethics approval will not be obtained for this review of published and publicly accessible data. Our results will be disseminated through conference presentations and a peer-reviewed publication. Our findings will inform implementation strategies that support the adoption of chiropractic care within integrated health care delivery systems.

**Strengths and limitations of this study:**

- This scoping review will be among the first to comprehensively map literature of doctors of chiropractic (DCs) working with or within military, veteran or civil integrated health care delivery systems.
- The literature search strategy is comprehensive and potentially generalizable to a global DC workforce, and relevant to other nonpharmacologic therapy providers who typically work in the community, e.g., acupuncturists, psychologists.
- A multidisciplinary team with diverse clinical and research expertise will inform our scoping review across all stages of the work.
- We have organized our search strategy and extraction form/guide around standardized terminology from the field of implementation science. It may be challenging to identify relevant studies, or extract all useful information, if original research does not use this terminology.
- Non-English articles describing the implementation of chiropractic care in a medical setting may be missed.

## INTRODUCTION

Musculoskeletal conditions, including back and neck pain, are leading causes of disability worldwide.^1^ In the United States, the use of pharmacologic treatments, such as opioids, and invasive procedures, such as steroid injections and surgery, for low back pain, increased from 1997 to 2010.^2^ During the same time period disability and costs from low back pain also increased.^2,3^ In contrast to these patterns of care for spinal disorders, clinical practice guidelines emphasize the use of nonpharmacologic approaches before the use of over the counter medications, prescribed medications, or invasive procedures.^4-7^ Yet patients who seek care in integrated health care delivery systems, at specific medical settings such as primary care clinics in hospitals or community health centers, still frequently receive prescribed medications as first line care.^8,9^ Limited familiarity with the efficacy and role of nonpharmacologic treatments, few opportunities to practice in the same location as nonpharmacologic providers, and inadequate channels of communication between these providers have been identified as important clinician-level barriers that prevent referrals to nonpharmacologic treatments.^10,11^ Increasing collaboration between primary care providers and providers of nonpharmacologic treatment will improve access to nonpharmacologic treatments and may improve outcomes.

Doctors of chiropractic (DCs) are trained to manage common spinal disorders, and can provide care that is aligned with international clinical practice guidelines for low back pain, lumbar spinal stenosis, neck pain, and headache.^4-7,12-15^ Chiropractic care is typically comprised of patient evaluation and evidence-based management, including patient education, exercise instruction, spinal manipulation or mobilization, and soft tissue therapy.^16,17^ Interestingly, patients who access chiropractic care for their low back pain are less likely than those who initiate care with a primary care provider to receive an opioid medication within one year, adjusting for sociodemographic characteristics and number of chronic conditions.^8,18^

Although DCs practice in at least 90 countries, chiropractic care is often unavailable in integrated health care delivery systems.^19^ The majority of DCs practice in private clinics based in the middle- or upper-income communities^20,21^ of high-income countries.^19^ Limited access to chiropractic care may be most pronounced in low-income settings where out-of-pocket costs and a scarcity of community-based DCs may limit use.^22^ While primary care providers may encourage their patients to seek chiropractic care, limited exposure to DCs and poor channels of communication may stifle such collaboration.^11^ Improved collaboration between medical providers and community-based DCs, or embedding DCs in medical settings, may improve access to evidence-based care for musculoskeletal conditions.

Successful expansion of chiropractic services in the Veterans Health Administration, the largest integrated health care delivery system in the United States, has demonstrated that providing chiropractic care within medical settings is feasible.^23^ Systematic reviews have also described the delivery of chiropractic care in active duty military settings.^24-26^ These reviews have focused on the characteristics of DCs practicing in military settings, common conditions managed by DCs, and clinical outcomes observed. A recent review evaluated implementation strategies used to increase access to musculoskeletal care within military settings, but not chiropractic care specifically.^28^ Review authors found that implementation strategies have focused on developing collaborative models of care, supporting communication between providers and hosting educational meetings. The review also highlighted the need to understand better the steps and details of the implementation process considering their scant description in the included studies. This gap in knowledge highlights the need to further develop and evaluate implementation strategies, defined as the methods to enhance the adoption, implementation, sustainment, and scale-up of an evidence-based practice.^27^ Furthermore, literature on the provision of chiropractic care in civil medical settings has not been systematically evaluated. Identifying strategies for implementing chiropractic care in low-income settings is particularly needed.

This scoping review aims to map studies of DCs working with or within medical settings, to identify implementation strategies used, and to describe the associated implementation and clinical outcomes. While some barriers to accessing chiropractic care may be unique, we anticipate our findings will be relevant to other providers of nonpharmacologic treatments not typically available in medical settings (e.g., acupuncturists, psychologists, and others). This scoping review is an important initial step in developing novel and testable implementation strategies that may increase access to nonpharmacologic treatments in primary care and other medical settings and decrease the overreliance on pharmacologic-based therapies such as opioids.

## METHODS

### Overview

Scoping reviews are a flexible and comprehensive methodology that examine the amount, variety, and characteristics of a broad research question.^29^ Our review will map literature of DCs working with or within integrated health care delivery systems, an emerging area of research. Thus, a scoping review, rather than a systematic review, is appropriate given the relatively unexplored nature of our research question.^30^ Our scoping review protocol follows methods recommended by Arksey^31^ and Levac^32^ which have since been formalized by the Joanna Briggs Institute^33^ and recent PRISMA-ScR guidelines^29^. Our approach involves the following six steps: 1) choosing the research question; 2) identifying relevant studies; 3) study selection; 4) charting or extracting the data; 5) collating, summarizing, and reporting the results; and 6) consultation with experts and stakeholders.

### Research question

Our study questions are guided by the Proctor Conceptual Model for Implementation Research.^34^ The Proctor Conceptual Model identifies three important and distinct concepts in implementation research: the clinical intervention, implementation strategies, and outcome measures. Our overarching research question and three subquestions, which map to the Proctor Conceptual model, are provided below. These are further described in the **Online Supplementary Appendix 1** and illustrated in **Figure 1**.

**Figure 1.**
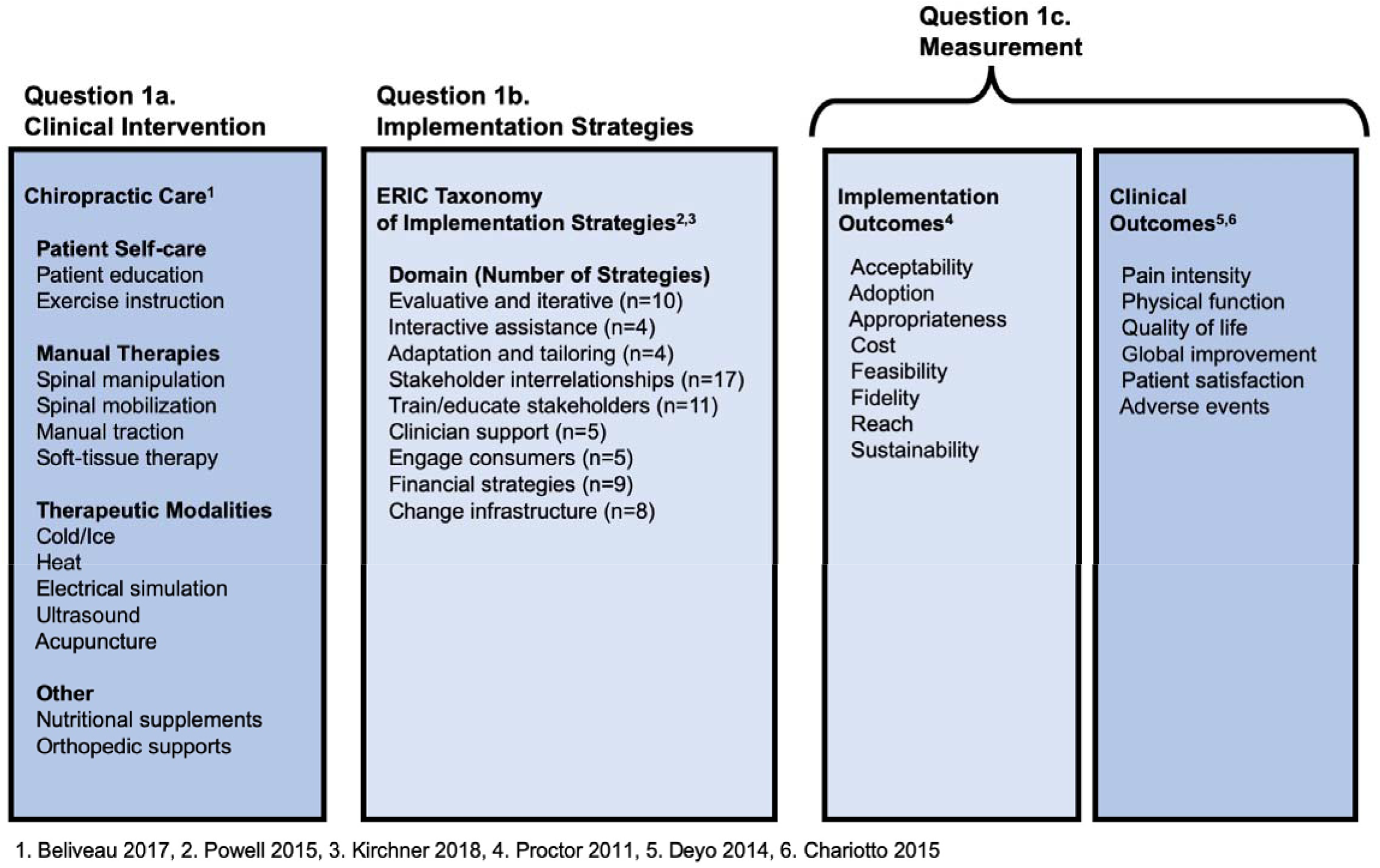
Key Concepts for Scoping Review Organized by Study Question and the Three Domains of the Proctor Conceptual Model for Implementation Research

**Question 1:** “What is known from existing peer-reviewed literature about DCs working with or within integrated health care delivery systems?”
  **Question #1a:** What are the characteristics of chiropractic care (e.g., location of service, department affiliation, conditions managed, treatments provided) in collaborative models of care that include DC and medical providers?
  **Question #1b:** What implementation strategies have been used to initiate or enhance these collaborative models?
  **Question #1c:** What implementation and clinical outcomes have been studied?

### Stage 2: Identifying relevant studies

We will identify English-language studies from 1998 to 2020 in three databases: MEDLINE, EMBASE and Web of Science. The cited references of included studies will be hand searched to identify additional potentially relevant studies. Our preliminary search strategy was developed with the assistance of the head librarian at the Boston University Medical Library (**Online Supplementary Appendix 2**). Relevant citations will be uploaded into Endnote, version X8.1 (Clarivate Analytics, Philadelphia, PA).

### Stage 3: Study selection

Our scoping review will include original studies that report data on DCs who work with or within medical settings. Our final eligibility criteria will be developed post hoc through an iterative team-based approach of reviewing the literature. We will consider concepts used in implementation research, which is defined as the development and evaluation of strategies to integrate evidence-based interventions within specific clinical settings.^35^ All members of the study team will complete an open-source course developed by the National Institutes of Health (USA) comprised of six modules presenting foundational concepts in implementation research.^36^

#### Inclusion and exclusion criteria

Our preliminary set of inclusion criteria is as follows: We will include peer-reviewed original research (i.e., experimental, observational, and qualitative studies) and knowledge syntheses (i.e., systematic reviews) that: (1) describe the practice or initiation of formal collaborative relationships between medical settings and community-based DCs, e.g., shared credentialing, continuing education, or electronic health record; or (2) describe the practice or implementation of chiropractic care within medical settings, e.g., hospitals, community health centers; and (3) contribute information to one or more of the three domains of the Proctor Conceptual Model, i.e. clinical intervention, implementation strategy, or outcome measurement.^34^

Examples of medical settings include outpatient hospital, clinic, or other practice sites where providers include allopathic and osteopathic physicians, nurse practitioners, or physician assistants. As the provision of health care has become more complex and multidisciplinary, fewer clinicians are working in traditional independent solo or small group private practices. As a result, the medical setting of interest for this review broadly encompasses military, veteran, and civil integrated health care delivery systems. This may include closed model networks such as the Veterans Health Administration and Kaiser Permanente or hospital affiliated networks that include outpatient services such as Partners Health Care (now named Mass General Brigham) in eastern Massachusetts and Intermountain Healthcare in the Intermountain West of the United States. Common to all of these delivery systems is the provision of a wide range of health care services within the provider network.

We will exclude non-English language studies and publications that do not describe original research (e.g., guidelines, editorials, commentaries). We will exclude studies that do not involve a medical setting (e.g., effectiveness trials based in chiropractic clinics only), as well as studies focusing on medical settings but without the provision of chiropractic care within a collaborative model of care.

All titles, abstracts, and relevant full-text articles will be screened independently by two reviewers using the final eligibility criteria. A third reviewer will evaluate conflicts between reviewers. If consensus cannot be reached, a senior investigator will be consulted. The use of a third reviewer and senior investigator will help ensure that appropriate application of eligibility criteria is utilized and all discrepancies are resolved. The screening process will be facilitated by Covidence software (Veritas Health Innovation, Melbourne, Australia).

### Stage 4: Charting and extracting data

A preliminary extraction form and guide developed by our multidisciplinary team are presented in **Online Supplementary Appendices 3 and 4**, respectively. Two team members will extract information from each study, independently, using this form. This will be an iterative process, and the extraction form will be updated as necessary. The purpose of this process is to collect information that can be mapped back to the Proctor Conceptual Model as illustrated in **Figure 1**. We anticipate collecting information from the following four categories: study characteristics, chiropractic care, implementation strategies and outcome measurement. When this information is incomplete or missing from published reports, we will contact authors of eligible manuscripts to request additional documentation (e.g., unpublished reports, protocols).

#### Study characteristics

We will collect information on study characteristics, e.g., study title, authors, country, year published, and journal. We will characterize the journal type, e.g., chiropractic, complementary and integrative health, general medicine, physical medicine and rehabilitation. We will identify the study design and whether a theory or framework related to implementation research or another field was used. We will indicate which stakeholders were involved in data collection (patients, clinicians, other) and how many contributed data.

#### Chiropractic care

We will report the number of DCs described in each study and describe their characteristics (e.g., age, gender, years in practice), and the number of clinics represented in each manuscript. Each clinical or set of clinics will be characterized as serving military, veteran, or civil populations. If integration of DCs into multiple medical clinics is described we will indicate whether the clinics are independent of each other or part of the same integrated health care delivery system. We will extract a description of each clinical setting and characterize it as primary care (internal or family medicine), physical medicine and rehabilitation, complementary and integrative medicine, orthopedics, pain medicine, or other. We will also identify in each study the hospital or clinic department affiliation of chiropractors if applicable. We will extract Information on which conditions were commonly managed by DCs (e.g., back pain, neck pain, headache), along with the treatments delivered, including guidance on patient self-care (patient advice and education, exercise instruction), manual therapies (spinal manipulation, spinal mobilization, manual traction, soft-tissue therapy), acupuncture, therapeutic modalities (cold/ice, heat, electrical stimulation, ultrasound), and other (nutritional supplements, orthopedic supports). Extraction of information on conditions managed and treatments provided by DCs will include ICD and CPT codes when available.

#### Implementation strategies

Each study will be characterized as describing a formal or informal implementation strategy or no implementation strategy. The label of ‘formal’ implementation strategy will be applied to studies that use the term ‘implementation strategy’. Formal or informal implementation strategies will be labelled using the Expert Recommendations for Implementing Change (ERIC) taxonomy, which includes 9 domains with 73 distinct strategies.^37-39^

#### Outcome measurement

We will identify studies that present implementation and/or clinical outcomes. Implementation outcomes include acceptability, adoption, appropriateness, feasibility, fidelity, implementation cost, penetration, and sustainability.^40^ Clinical outcomes include pain intensity, physical function, health-related quality of life, global improvement, patient satisfaction, and adverse events.^41,42^

### Stage 5: Collating, summarizing and reporting the results

Summaries of findings from each extraction form will be entered into a matrix and emerging themes will be discussed at regular team meetings. We will provide a descriptive overview of the eligible individual studies and themes using tables and/or graphical summaries. Separate tables or graphical summaries well be used to organize information on each of the three domains of the Proctor Conceptual Model: description of chiropractic care, implementation strategies, and outcome measurement. Within each summary we will organized studies by setting (active military, veteran, civil) to help demonstrate potential similarities and differences that may impact the implementation process. Altogether, this organization of data characteristics and themes will facilitate the development of implementation strategies that can be tailored to particular needs of an individual practice or integrated health care delivery system. The summary material will form the basis for a report which will be organized according to PRISMA-ScR gudielines.^29^

### Stage 6: Consultation

The purpose of the consultation phase is to stimulate ongoing feedback and maximize the usefulness of our findings to providers from a range of healthcare disciplines that may collaborate with DCs in routine clinical care. Consultation will involve two modes. First, our core multidisciplinary research team will provide ongoing feedback across all stages of the review. Second, we will conduct a brief online survey with a larger group of stakeholders to generate feedback on our key findings and their implications.

As we developed this protocol we formed a multidisciplinary team that will continue to offer feedback throughout the conduct of our scoping review. Monthly team meetings and ad hoc document review will serve as a vehicle for ongoing feedback. Team members represent clinical expertise in primary care (internal and family medicine), pediatrics, geriatrics, physical medicine and rehabilitation, rheumatology, pain medicine, addiction medicine, chiropractic care, and complementary and integrative medicine.

We will conduct a brief cross-sectional online survey with a larger group of stakeholders who possess expertise and interest in the purpose of this scoping review. The purpose of this survey is to generate additional feedback on our findings from a broader group of participants. Each of the members of the research team will assist in recruiting members of their disciplines to participate in the survey resulting in a convenience sample of at least 30 participants who represent a diverse range of healthcare fields. Survey questions will be administered before and after participants read a brief report that summarizes our key findings. We anticipate including a mixture of closed and open-ended questions, although questions will be determined based on findings from scoping review and discussion at team meetings.

## ETHICS AND DISSEMINATION

Formal ethical approval is not required to undertake this scoping review of published and publicly accessible literature. The completed scoping review will be submitted for publication to a peer-reviewed, interdisciplinary open access journal, in addition to conferences attended by primary care providers, medical specialists, and chiropractors. Our findings will form the basis of implementation strategies to support adoption of chiropractic care within integrated health care delivery systems. Ultimately, we anticipate these strategies will improve access to a broader range of evidence-based nonpharmacologic treatments for common painful musculoskeletal conditions (e.g., acupuncturists, psychologists, others), which may reduce reliance on pharmacologic-based therapies such as opioids.

## Data Availability

This is a protocol paper.

## AUTHOR CONTRIBUTIONS

Study concept and design: Roseen, Kasali, Corcoran, Masselli, Laird, Saper, Alford, Cohen, Lisi, Atlas, Bean, Evans, Bussieres

Acquisition, analysis, or interpretation of data: n/a

Drafting of the manuscript: Roseen

Critical revision of the manuscript for important intellectual content: Roseen, Kasali, Corcoran, Masselli, Laird, Saper, Alford, Cohen, Lisi, Atlas, Bean, Evans, Bussieres

Statistical analysis: n/a

Obtained funding: n/a

Administrative, technical, or material support: Roseen, Kasali, Corcoran, Masselli Study supervision: Roseen, Saper, Laird, Evans, Bussieres

## Appendices

**Online Supplementary Appendix 1:** Description of conceptual framework

**Online Supplementary Appendix 2:** Search strategy

**Online Supplementary Appendix 3:** Extraction form

**Online Supplementary Appendix 4:** Extraction guide

### Online Supplementary Appendix 1. Description of conceptual framework

**Implementation science** is “the scientific study of methods to promote the systematic uptake of research findings and other evidence-based practices into routine practice, and hence, to improve quality and effectiveness of health services”.^1^

The **Proctor Conceptual Model** guides the process of evaluating both implementation and clinical outcomes of a specific clinical intervention. The Proctor Conceptual Model for the study of Implementation Research comprises three domains: clinical intervention, implementation strategies and outcome measurement.^2^

**Clinical Interventions** are evidence-based practices. The clinical intervention of interest in this scoping review is chiropractic care, which includes evaluation and treatment. Common treatments provided by DCs for back and neck pain include joint manipulation, soft tissue therapy, patient education, and exercise instruction.^3,4^ These approaches align with clinical practice guidelines for back pain, neck pain, and headache.^5-12^

**Implementation strategies** are defined as methods to enhance the adoption, implementation, sustainment, and scale-up of an innovation.^13^ Implementation strategies address facilitators and/or barriers to successful implementation at one or more socioecological levels, such as policy, systems environment, organizational structure, individual providers, and consumers. Furthermore, implementation strategies influence the uptake of the ‘clinical intervention’ through addressing barriers at one or more levels of influence (e.g. patient-, provider-, clinic-, and health-system factors) to facilitate implementation The Expert Recommendations for implementing Change (ERIC) Project taxonomy is a comprehensive organized list of implementation strategies.^14-16^ The ERIC Project encompasses a set of 73 implementation strategies across 9 domains.

**Outcome measurement**, in the Proctor Conceptual Model, differentiates implementation and clinical outcomes. Successful implementation is thought to be necessary for clinically efficacious interventions to be effective in real world settings.^2^ The Proctor Conceptual Model identifies and defines eight core implementation outcomes.^17^ Three of these implementation outcomes (acceptability, appropriateness, feasibility) are strong predictors of implementation success, sometimes considered pre-implementation outcomes. The other five (reach, adoption, fidelity, cost, penetration, and sustainability) are typically used to evaluate effectiveness of implementation strategies. Important clinical outcomes for adults with common musculoskeletal conditions include pain, physical function, quality of life, and patient satisfaction.^18,19^

**Online Supplementary Appendix 2:** Preliminary search strategy

|  |  |
| --- | --- |
| **Pubmed** | (((((("Referral and Consultation"[Mesh] OR "Interprofessional Relations"[Mesh] OR Implementation[Title/Abstract] OR Implement[Title/Abstract] OR Implemented[Title/Abstract] OR Integration[Title/Abstract] OR Integrated[Title/Abstract] OR Integrate[Title/Abstract] OR Dissemination[Title/Abstract] OR Disseminate[Title/Abstract] OR Disseminated[Title/Abstract] OR Roll out[Title/Abstract] OR Knowledge translation[Title/Abstract] OR Knowledge transfer[Title/Abstract] OR interdisciplinary[Title/Abstract] OR multidisciplinary[Title/Abstract])) AND (("Chiropractic"[Mesh] OR "Manipulation, Chiropractic"[Mesh] OR "Manipulation, Spinal"[Mesh] OR Chiropractic[Title/Abstract] OR Chiropractor[Title/Abstract] OR Spinal manipulation[Title/Abstract] OR Manual therapy[Title/Abstract] OR Non-pharmacologic[Title/Abstract] OR Spine care[Title/Abstract]))) AND ("Hospitals"[Mesh] OR "Delivery of Health Care"[Mesh] OR "Academic Medical Centers"[Mesh] OR "Health Facilities"[Mesh] OR Health care[Title/Abstract] OR Health care service[Title/Abstract] OR Healthcare[Title/Abstract] OR Healthcare service[Title/Abstract] OR Hospital[Title/Abstract] OR Health services research[Title/Abstract] OR Primary care[Title/Abstract] OR Community health center[Title/Abstract] OR Veteran's affairs[Title/Abstract] OR Veterans affairs[Title/Abstract] OR Utilization[Title/Abstract] OR Delivery[Title/Abstract] OR Service[Title/Abstract] OR Services[Title/Abstract] OR Hospital program[Title/Abstract] OR Hospital programs[Title/Abstract] OR Tertiary service[Title/Abstract] OR Tertiary services[Title/Abstract] OR medical center[Title/Abstract])) AND ( "1998/01/01"[Pdat] : "3000/12/31"[Pdat] ) AND English[lang])) |
| **Web of Science** | TS=(refer* OR consult* OR interprofessional relation* OR implement* OR integrat* OR disseminat* OR roll out OR knowledge trans* OR multidisciplin* OR interdisciplin*)<br>AND<br>TS=(chiroprac* OR spin* manipul* OR "manual therap*" OR non-pharmacologic OR spine care)<br>AND<br>TS=(hospital OR hospitals OR health center* OR health facilit* OR primary care OR community health center* OR veterans affairs OR veteran's affairs OR medical center*) |
| **Embase** | ('patient referral'/exp OR 'public relations'/exp OR 'implementation'/exp OR 'integration'/exp OR 'dissemination'/exp<br>OR 'knowledge translation'/exp OR 'knowledge transfer'/exp OR 'implement*':ab,ti OR 'integrat*':ab,ti OR<br>'disseminat*':ab,ti OR 'roll out':ab,ti OR 'knowledge translat*':ab,ti OR 'knowledge transfer*':ab,ti OR<br>'multidisciplin*':ab,ti OR 'interdisciplin*':ab,ti<br>AND<br>'chiropractic'/exp OR 'chiropractor'/exp OR 'chiropractic practice'/exp OR 'chiropractic manipulation'/exp OR 'spine<br>manipulation'/exp OR 'chiropract*':ab,ti OR 'spinal manipul*':ab,ti OR 'manual therapy':ab,ti OR 'non-<br>pharmacologic':ab,ti OR 'spine care':ab,ti<br>AND<br>'health care facility'/exp OR 'primary medical care'/exp OR 'veterans affairs hospital'/exp OR 'hospital':ab,ti OR<br>'health services research':ab,ti OR 'primary care':ab,ti OR 'community health center':ab,ti OR 'veterans affairs':ab,ti<br>OR 'hospital program*':ab,ti OR 'medical center*':ab,ti) |

**Online Supplementary Appendix 3.** Extraction form

| Reviewer: |  |  |
| --- | --- | --- |
| Date: |  |  |
| **Study characteristics** |  |  |
| Variable | Category | Notes / Excerpts from manuscript (page number) |
| Study title | ☐ _____<br>(please specify) |  |
| First author | ☐ _____<br>(please specify) |  |
| Publication year | ☐ _____<br>(please specify) |  |
| Study country | ☐ _____<br>(please specify) |  |
| Journal | Journal name _____<br>(please specify)<br>Select category<br>☐ Chiropractic<br>☐ Complementary and integrative health<br>☐ General medicine<br>☐ Physical medicine and rehabilitation |  |
|  | ☐ Other<br>(please specify: _____) |  |
| Number of medical settings represented | Name of integrated health care delivery system:<br>_____<br>(please specify)<br>How many clinics in study? _____<br>(please specify)<br>If multiple, are they:<br>☐ Independent<br>☐ Part of same healthcare system<br>☐ Unclear<br>(please specify: _____) |  |
| Departmental affiliation of medical setting(s) | ☐ Internal medicine<br>☐ Family medicine<br>☐ Physical medicine/rehabilitation<br>☐ Integrative medicine<br>☐ Orthopedics<br>☐ Pain medicine<br>☐ Other<br>(please specify: _____)<br>☐ Not applicable |  |
| Characteristics of DCs | ☐ Number of DCs<br>(please specify: _____)<br>☐ Average age<br>(please specify: _____)<br>☐ Gender<br>(please specify: _____ % female)<br>☐ Average Years in practice<br>☐ Department affiliation<br>(please specify: _____) |  |
| Departmental affiliation of medical setting | ☐ Internal medicine<br>☐ Family medicine<br>☐ Physical medicine/rehabilitation<br>☐ Integrative medicine<br>☐ Orthopedics<br>☐ Pain medicine<br>☐ Other<br>(please specify: _____)<br>☐ Not applicable |  |
| Study design | ☐ Systematic review<br>☐ Observational/Cross-sectional<br>☐ Observational/Longitudinal<br>☐ Qualitative<br>☐ Mixed-methods<br>☐ Randomized control trial<br>☐ Other<br>(please specify: _____) |  |
| Description of patient population | Indicate the proportion of patients seen for the following conditions:<br>☐ Back pain<br>(please specify: _____%)<br>☐ Neck pain<br>(please specify: _____%)<br>☐ Headache<br>(please specify: _____%)<br>☐ Other<br>(please specify: _____%)<br>☐ Not Reported |  |
| **Clinical intervention: Chiropractic care** |  |  |
| Patient Self-care | ☐ Patient Education<br>☐ Exercise Instruction |  |
| Manual Therapies | ☐ Spinal Manipulation<br>☐ Spinal Mobilization<br>☐ Manual Traction<br>☐ Soft-tissue Therapy |  |
| Therapeutic Modalities | ☐ Cold/ice<br>☐ Heat<br>☐ Electrical Simulation<br>☐ Ultrasound<br>☐ Acupuncture |  |
| Other | ☐ Nutritional Supplements<br>☐ Orthopedic Supports |  |

| Implementation strategies |  |
| --- | --- |
| Description on implementation strategies | Which best characterizes the description of implementation strategies in manuscript?<br>☐ No description<br>☐ Informal<br>☐ Formal |
| ERIC taxonomy | If formal/informal description of implementation strategies is available, which of the 79 implementation strategies listed below were described? |
| Evaluative and iterative strategies | ☐ Assess for readiness and identify barriers and facilitators<br>☐ Audit and provide feedback<br>☐ Conduct cyclical small tests of change<br>☐ Conduct local needs assessment<br>☐ Develop a formal implementation blueprint<br>☐ Develop and implement tools for quality monitoring<br>☐ Develop and organize quality monitoring systems<br>☐ Obtain and use patient s/consumers and family feedback<br>☐ Purposefully reexamine the implementation<br>☐ Stage implementation scale up |
| Interactive assistance | ☐ Centralize technical assistance<br>☐ Facilitation<br>☐ Provide clinical supervision<br>☐ Provide local technical assistance |
| Adapt and tailor to context | ☐ Promote adaptability<br>☐ Tailor strategies<br>☐ Use data experts<br>☐ Use data warehousing techniques |
| Develop stakeholder interrelationships | ☐ Build a coalition<br>☐ Capture and share local knowledge<br>☐ Conduct local consensus discussions<br>☐ Develop academic partnerships<br>☐ Develop an implementation glossary<br>☐ Identify and prepare champions<br>☐ Identify early adopters<br>☐ Inform local opinion leaders<br>☐ Involve executive boards<br>☐ Model and stimulate change<br>☐ Obtain formal commitments<br>☐ Organize clinician implementation team meetings<br>☐ Promote network weaving<br>☐ Recruit, designate, and train for leadership<br>☐ Use advisory boards and workgroups<br>☐ Use an implementation advisor<br>☐ Visit other sites |
| Train and educate stakeholders | ☐ Conduct educational meetings<br>☐ Conduct educational outreach visits<br>☐ Conduct ongoing training<br>☐ Create a learning collaborative<br>☐ Develop educational materials<br>☐ Distribute educational materials<br>☐ Make training dynamic<br>☐ Provide ongoing consultation<br>☐ Shadow other experts<br>☐ Use train-the-trainer strategies<br>☐ Work with educational institutions |
| Support clinicians | ☐ Create new clinical teams<br>☐ Develop resource sharing agreements<br>☐ Facilitate relay of clinical data to providers<br>☐ Remind clinicians<br>☐ Revise professional roles |
| Engage consumers | ☐ Increase demand<br>☐ Intervene with patients/consumers to enhance uptake and adherence<br>☐ Involve patients/consumers and family members<br>☐ Prepare patients/consumers to be active participants<br>☐ Use mass media |
| Utilize financial strategies | ☐ Access new funding<br>☐ Alter incentive/allowance structures<br>☐ Alter patient/consumer fees<br>☐ Develop disincentives<br>☐ Fund and contract for the clinical innovation<br>☐ Make billing easier<br>☐ Place innovation on fee for service lists/formularies<br>☐ Use capitated payments<br>☐ Use other payment schemes |
| Change infrastructure | ☐ Change accreditation or membership requirements<br>☐ Change liability laws<br>☐ Change physical structure and equipment<br>☐ Change record systems<br>☐ Change service sites<br>☐ Create or change credentialing and/or licensure standards<br>☐ Mandate change<br>☐ Start a dissemination organization |

| Outcome measurement |  |
| --- | --- |
| Implementation outcomes | ☐ Acceptability<br>☐ Adoption<br>☐ Appropriateness<br>☐ Feasibility<br>☐ Fidelity<br>☐ Implementation Cost<br>☐ Penetration<br>☐ Sustainability |
| Clinical outcomes | ☐ Pain intensity<br>☐ Physical functioning<br>☐ Health-related quality of life<br>☐ Global improvement<br>☐ Patient satisfaction<br>☐ Adverse events |

**Online Supplementary Appendix 4.**
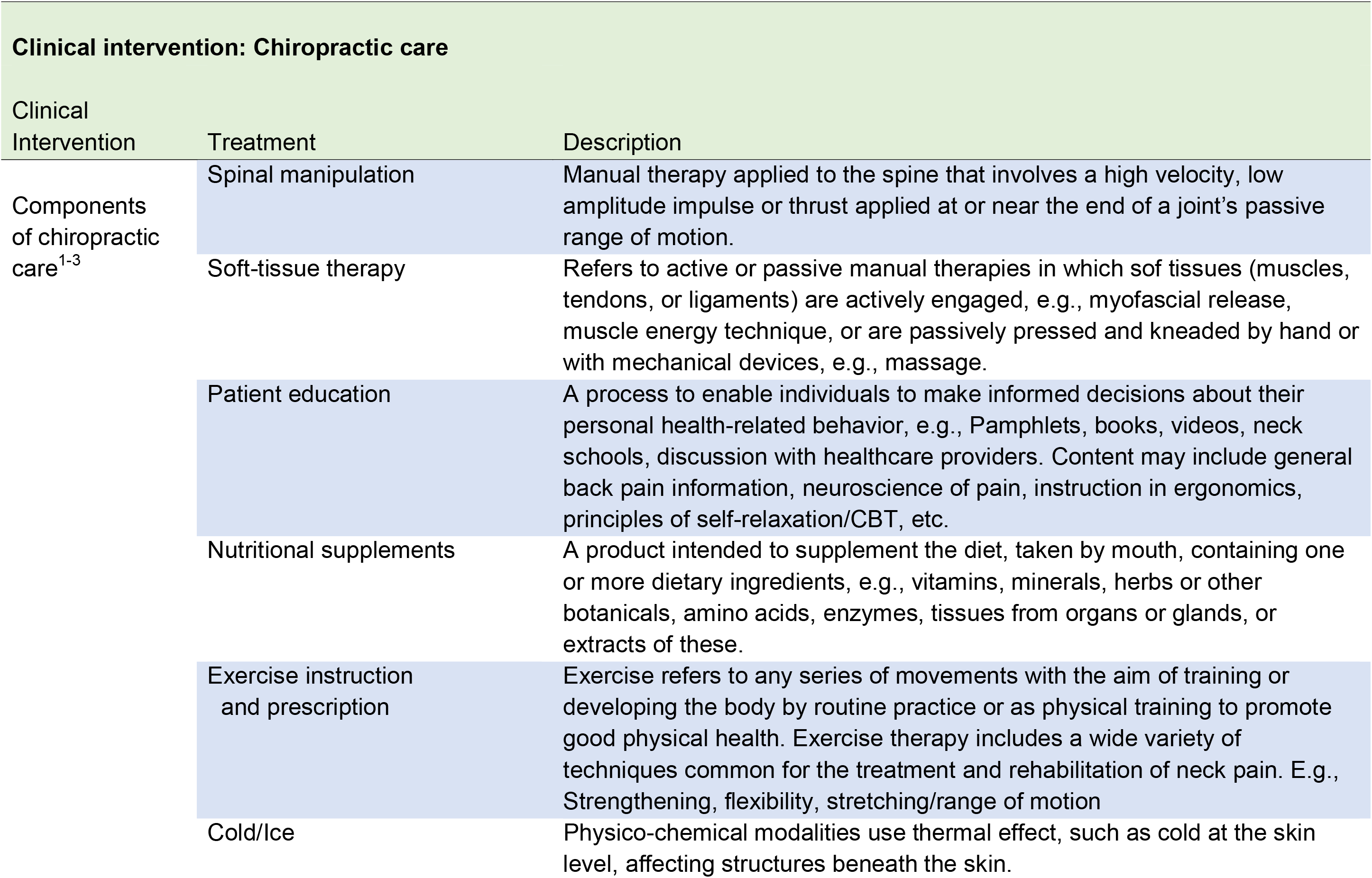
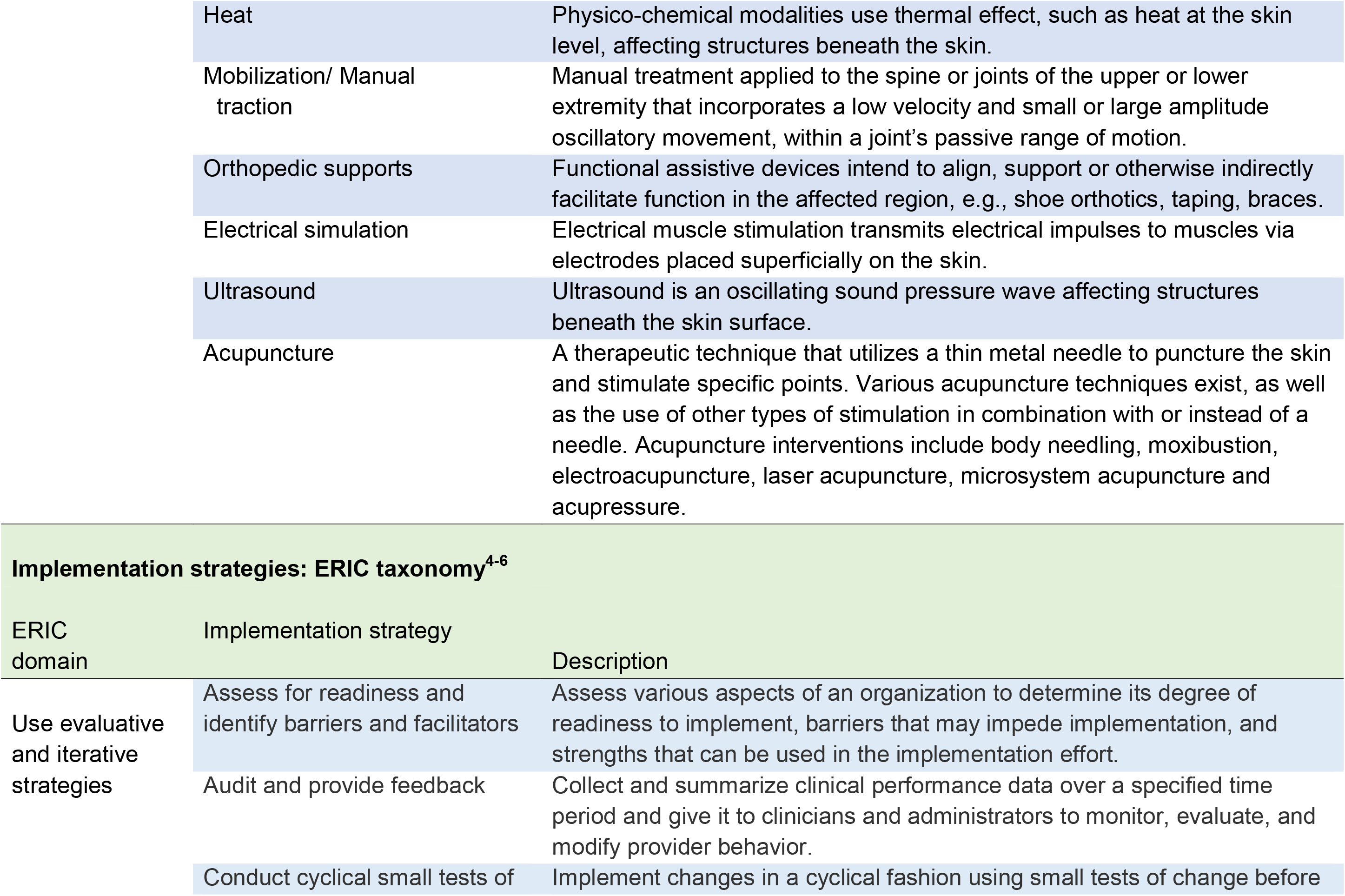
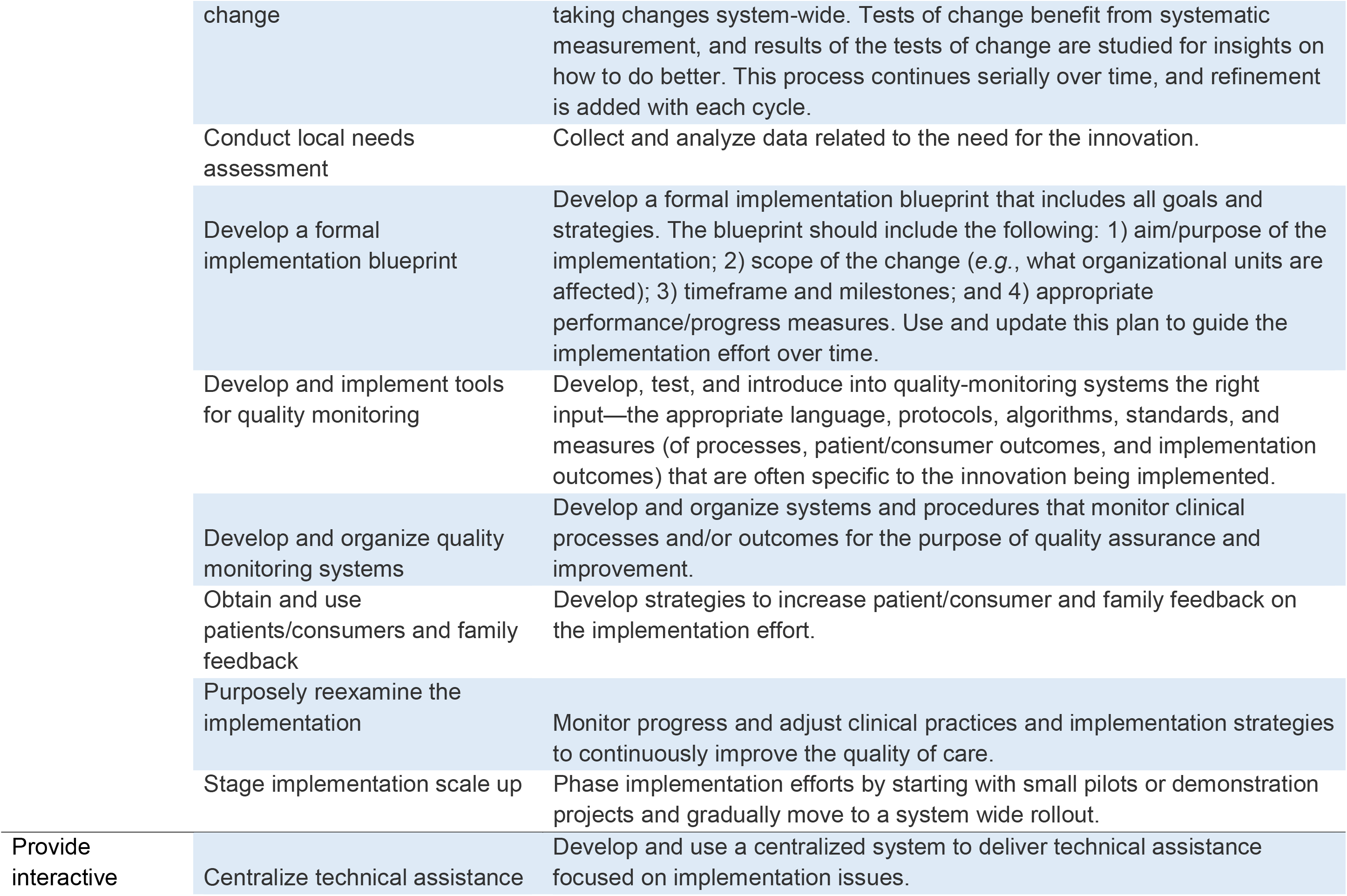
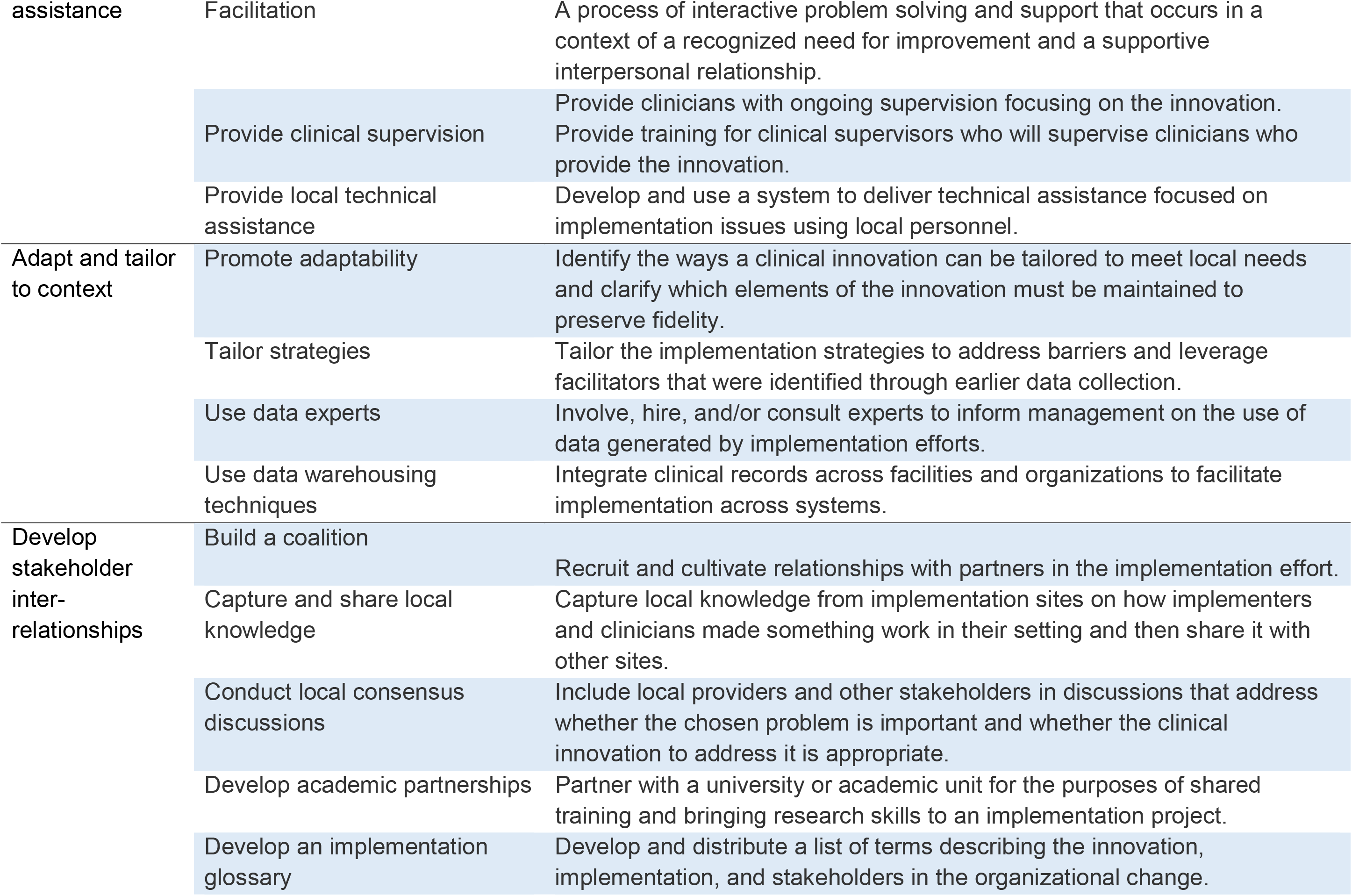
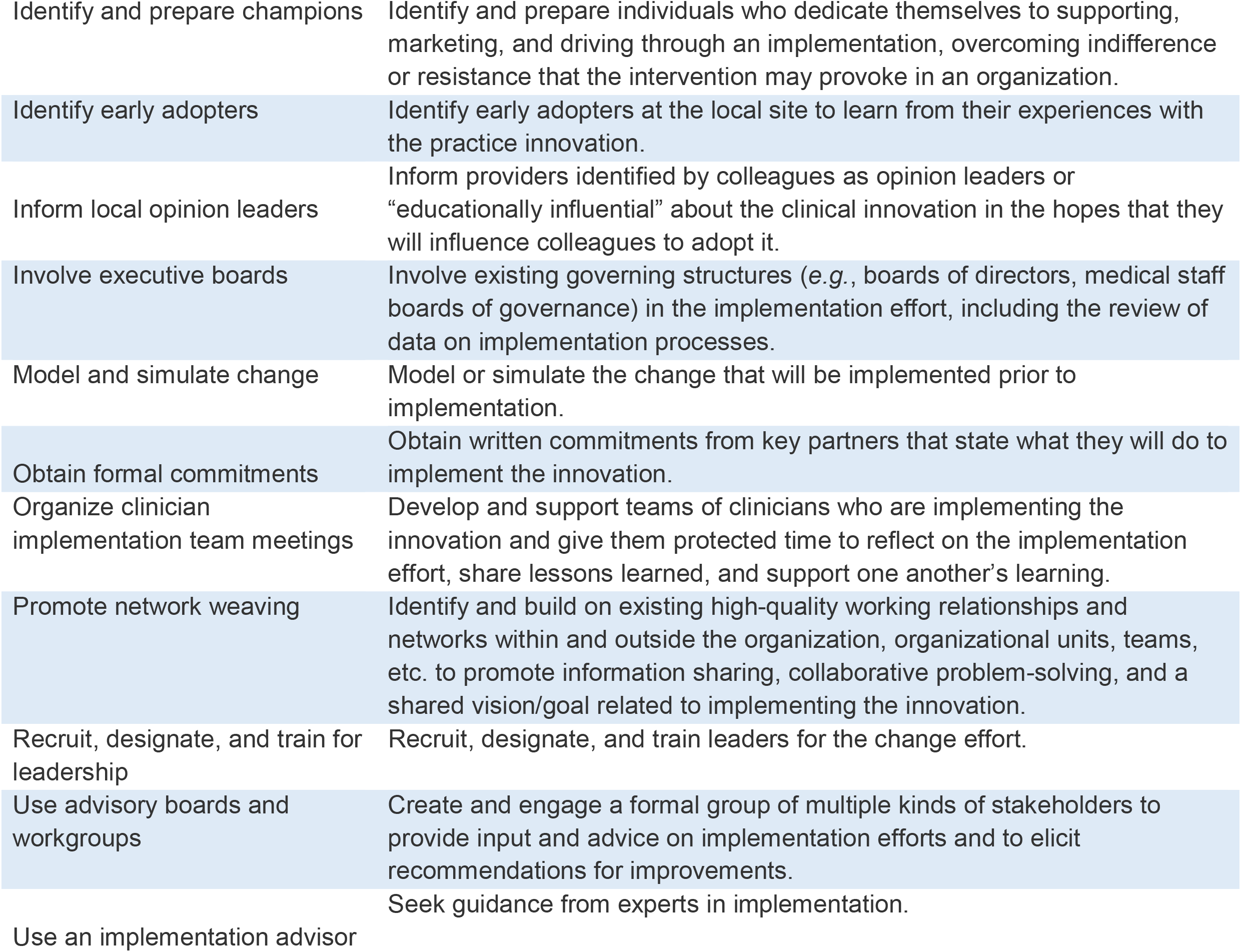
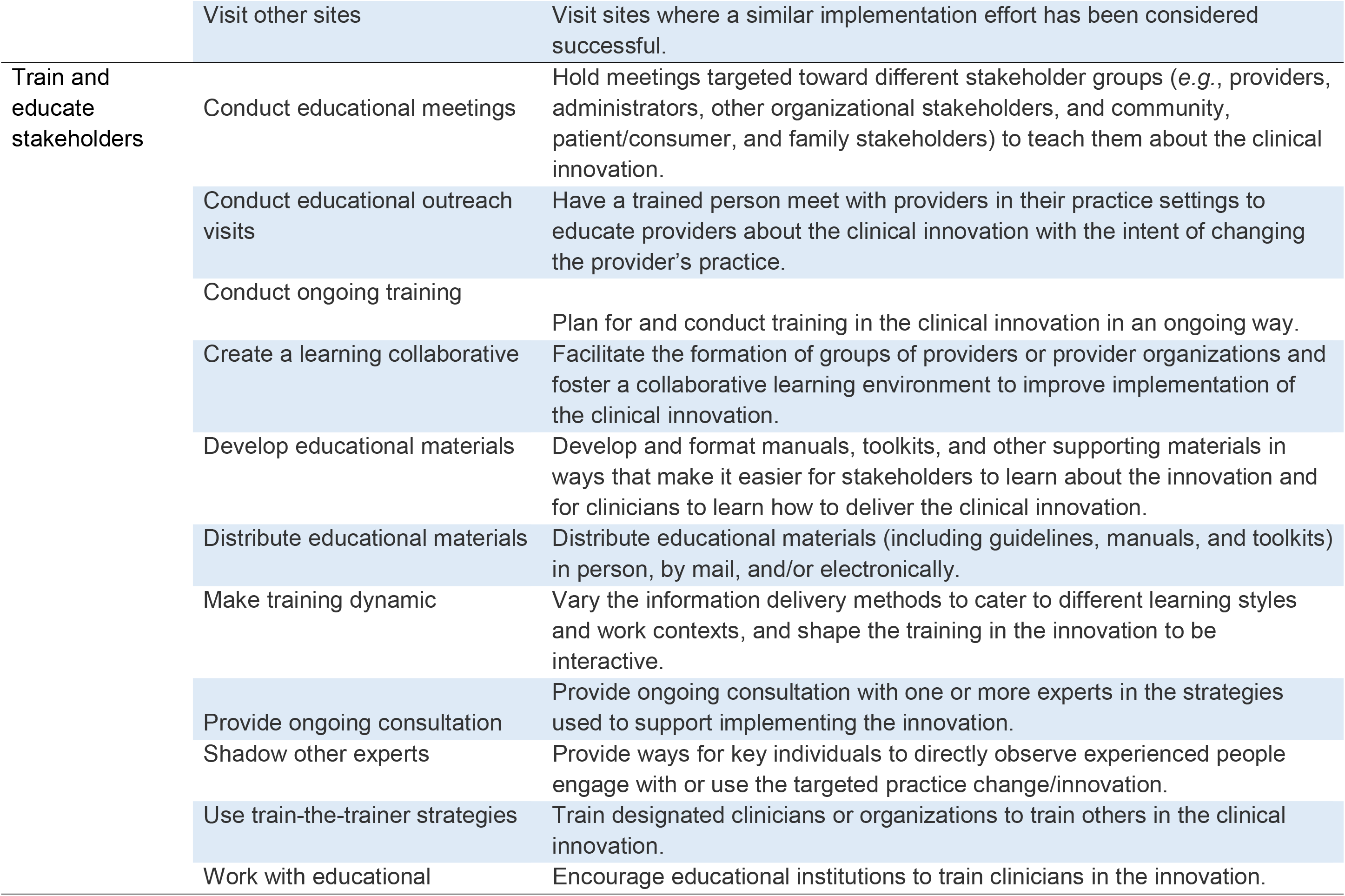
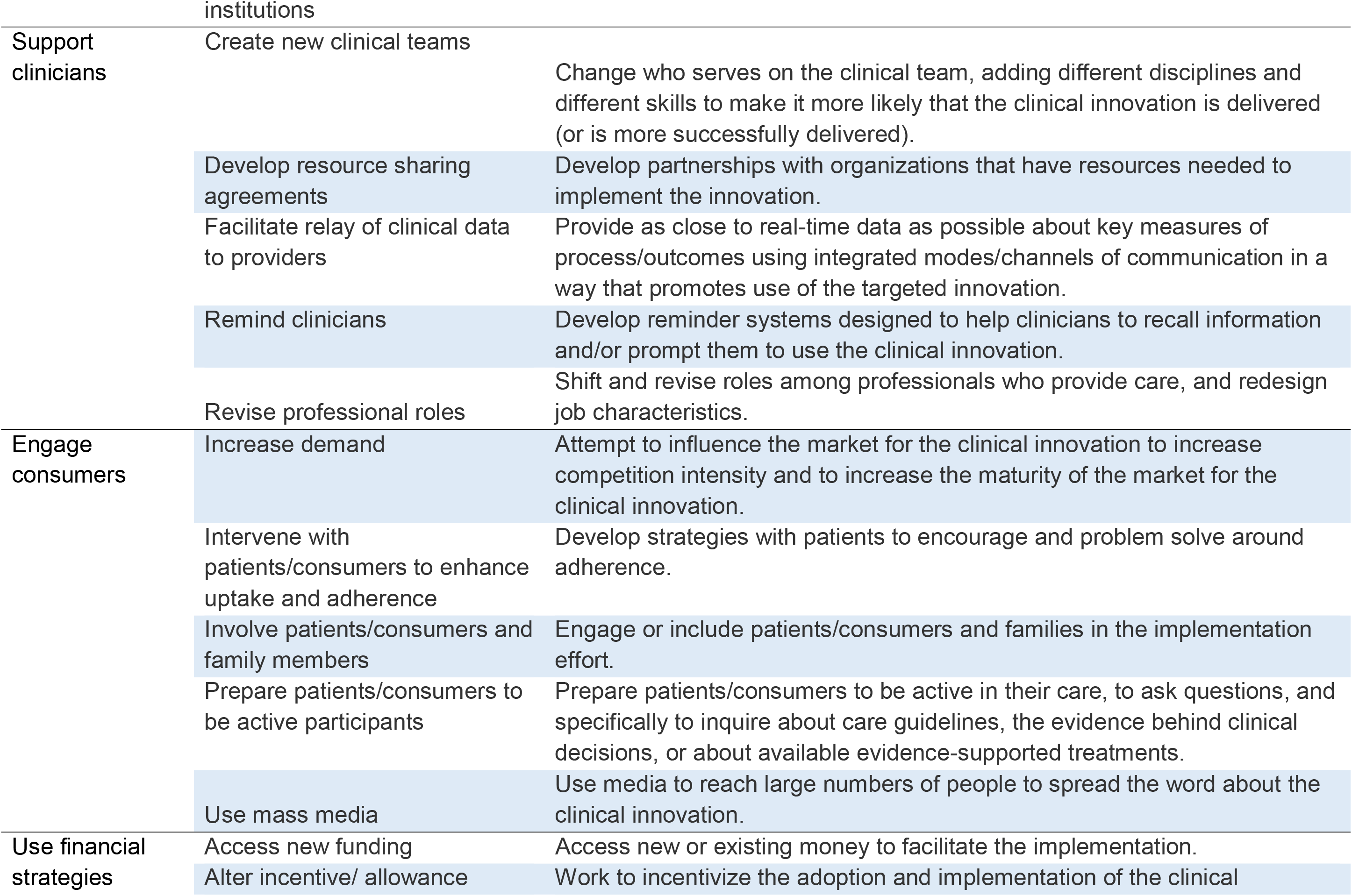
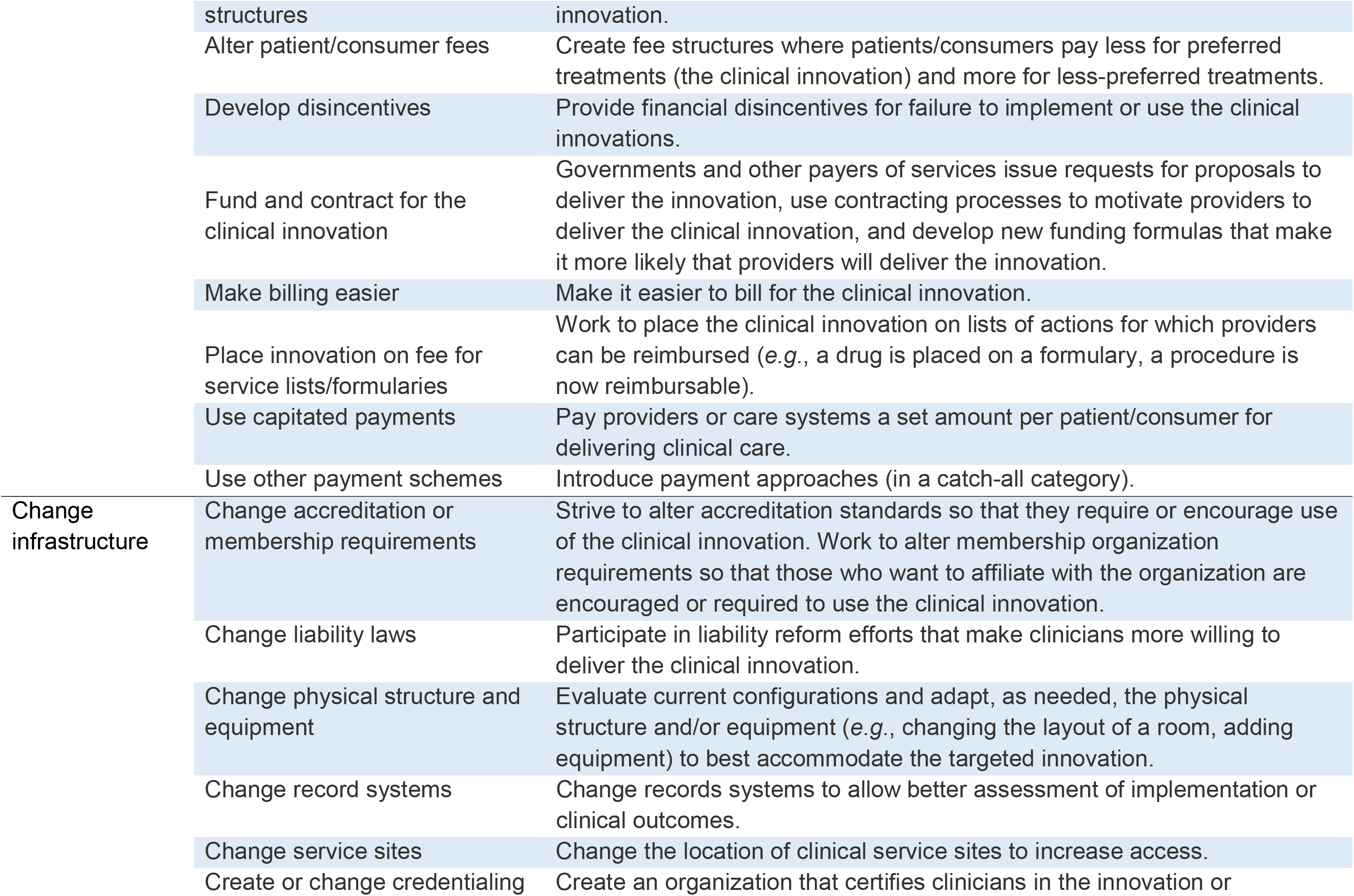
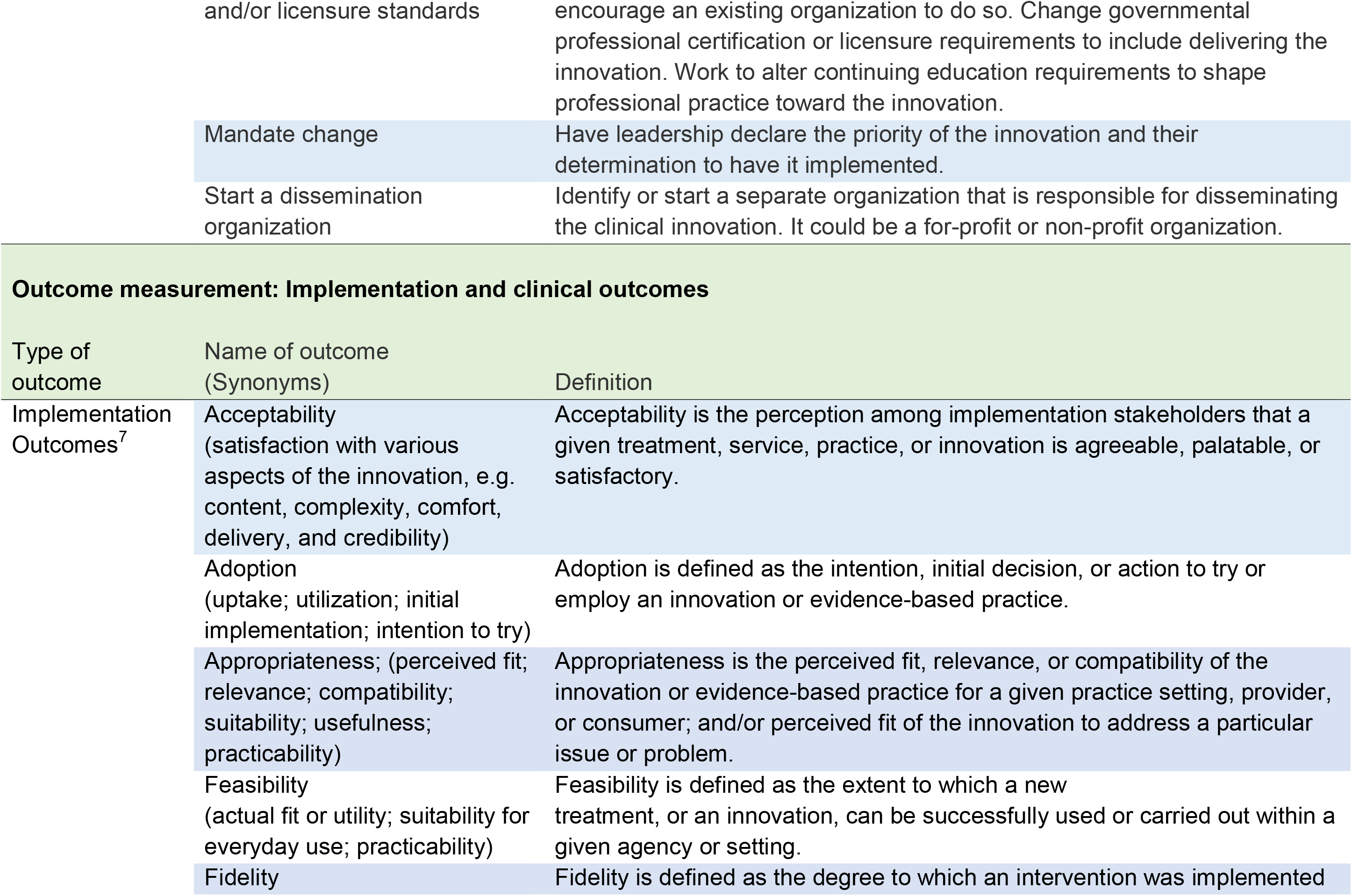
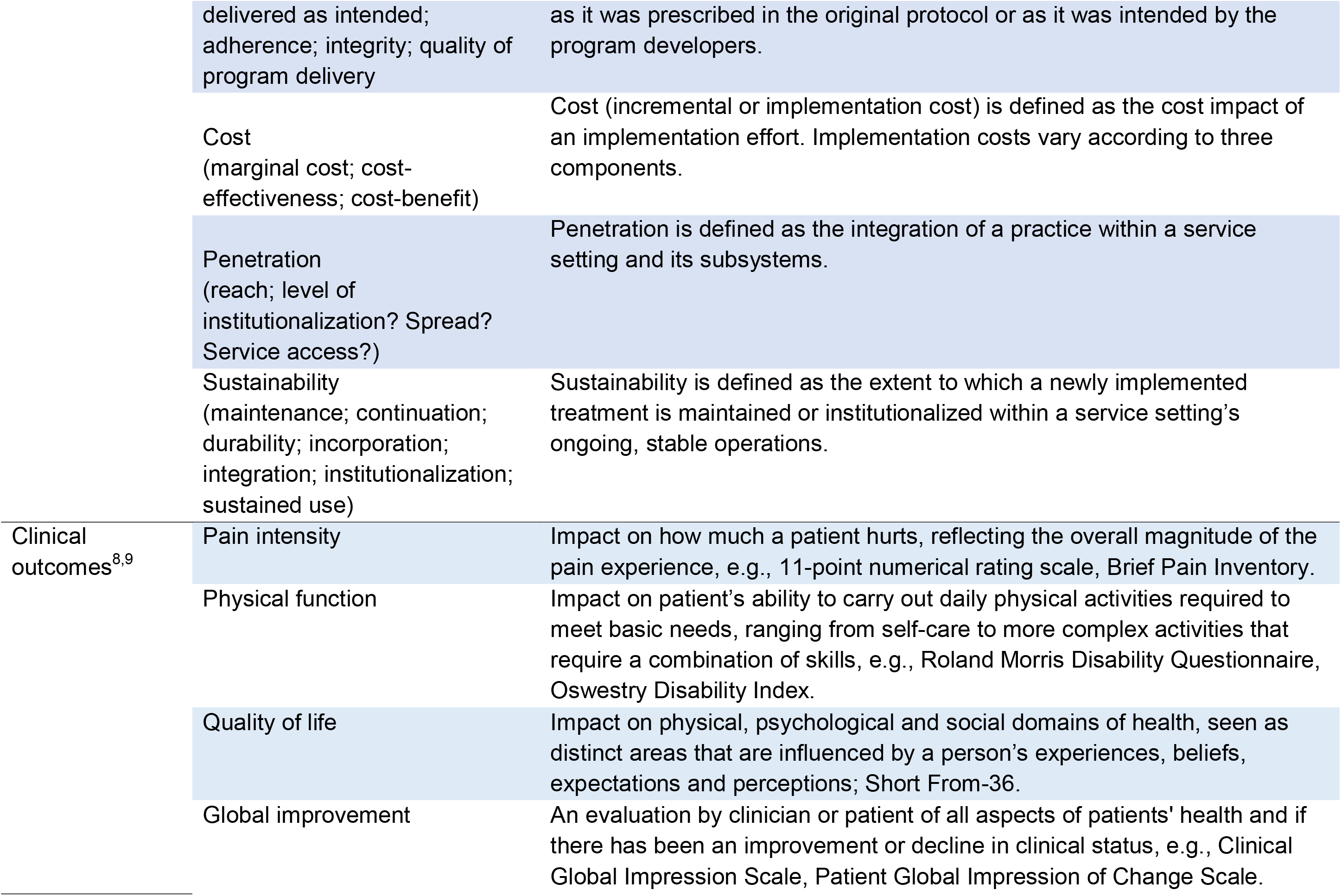
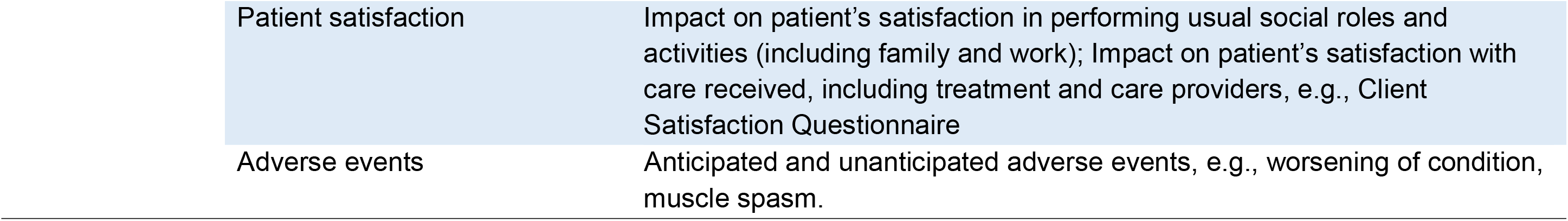
Extraction guide

